# Vaccine effectiveness and duration of protection of Comirnaty, Vaxzevria and Spikevax against mild and severe COVID-19 in the UK

**DOI:** 10.1101/2021.09.15.21263583

**Authors:** Nick Andrews, Elise Tessier, Julia Stowe, Charlotte Gower, Freja Kirsebom, Ruth Simmons, Eileen Gallagher, Meera Chand, Kevin Brown, Shamez N Ladhani, Mary Ramsay, Jamie Lopez Bernal

## Abstract

**Background:** COVID-19 vaccines have been used for 9 months in the UK. Real world data have demonstrated the vaccines to be highly effective against COVID-19, severe disease and death. Here, we estimate vaccine effectiveness over time since the second dose of Comirnaty, Vaxzevria and Spikevax in England.

**Methods:** We used a test-negative case-control design to estimate vaccine effectiveness against symptomatic disease, hospitalisation and mortality by age, comorbidity status and over time after the second dose to investigate waning separately for Alpha and Delta variants.

**Results:** Vaccine effectiveness against symptomatic disease peaked in the early weeks after the second dose and then fell to 47.3 (95% CI 45 to 49.6) and 69.7 (95% CI 68.7 to 70.5) by 20+ weeks against the Delta variant for Vaxzevria and Comirnaty, respectively. Waning of vaccine effectiveness was greater for 65+ year-olds compared to 40-64 year-olds. Vaccine effectiveness fell less against hospitalisations to 77.0 (70.3 to 82.3) and 92.7 (90.3 to 94.6) beyond 20 weeks post-vaccination and 78.7 (95% CI 52.7 to 90.4) and 90.4 (95% CI 85.1 to 93.8) against death for Vaxzevria and Comirnaty, respectively. Greater waning was observed among 65+ year-olds in a clinically extremely vulnerable group and 40-64-year olds with underlying medical conditions compared to healthy adults.

**Conclusions:** We observed limited waning in vaccine effectiveness against hospitalisation and death more than 20 weeks post-vaccination with Vaxzevria or Comirnaty. Waning was greater in older adults and those in a clinical risk group, suggesting that these individuals should be prioritised for booster doses.

## Background

Real world effectiveness data have consistently demonstrated high levels of short-term protection by COVID-19 vaccines against clinical disease and, more so, against severe outcomes including hospitalization and death.^1-7^ The duration of protection and, therefore, the need for booster doses, however, is uncertain.

Following the emergence of the Delta variant in the UK, we observed a slightly lower vaccine effectiveness against symptomatic disease following infection with the Delta variant compared to the Alpha variant in fully-vaccinated adults receiving an extended 12-week interval two-dose schedule for both the Pfizer-BioNTech Comirnaty (88.0% vs. 93.7%) and AstraZeneca Vaxzevria (67.0% vs 74.5%) vaccines. Other highly vaccinated countries have reported significantly reduced protection against infection following the emergence of the Delta variant. Among elderly nursing home residents in the United States where two doses of mRNA vaccines given 3 weeks apart was recently estimated to be 74.7% effective against symptomatic and asymptomatic SARS-CoV2 infection (from March to May 2021) but declined to 53.1% from June-July 2021, when the Delta variant was predominantly circulating.^8^ In Qatar, recent analysis observed no evidence of protection against infection from 20 weeks after vaccination. ^9^ Nevertheless, the extent to which reduced effectiveness is a result of the new variant or waning immunity is not yet clear. Immunogenicity data suggest that antibody titres wane relatively rapidly following two doses of vaccine, suggesting that waning may be an important factor in reported declines in vaccine effectiveness.^10^ The correlation between serological markers and protection against infection, disease or adverse outcomes, however, has not yet been established.

In the UK, COVID-19 vaccines have been in use since early December 2020. Initially a 3-week interval between doses of Comirnaty was used for approximately four weeks, but soon changed to an extended 12-week interval for all vaccines until June 2021 when the interval was reduced to 8 weeks following the emergence of the Delta variant. In this study, we estimate vaccine effectiveness over time since the second of Comirnaty, Vaxzevria, and Moderna Spikevax vaccines to investigate waning of protection against symptomatic disease, hospitalization and death separately for the Alpha and Delta variants.

## Methods

### Study Design

We used a test-negative case–control design to estimate vaccine effectiveness of two doses of Vaxzevria, Comirnaty and Spikevax COVID-19 vaccines against PCR-confirmed symptomatic disease, hospitalisation with 14 days of PCR confirmation, and death within 28 days of PCR confirmation. The analysis was stratified to asses vaccine effectiveness against the Delta and Alpha variants over the period that they were circulating, as described previously.^1^ In brief, we compared vaccination status in symptomatic adults with PCR-confirmed SARS-CoV-2 infection for each outcome of interest with vaccination status in adults who reported symptoms but had a negative SARS-CoV-2 PCR test. This approach helps to control for biases related to exposure, health-seeking behaviour, access to testing, and case ascertainment.

### Data sources

Sources of data on vaccination status, SARS-Cov-2 testing, identification of variants, covariates included in the analyses and data linkage methods have been described previously.^1^ Community testing data between 08 December 2020 and 03 September 2021 were included in the analysis. Data were restricted to persons who had reported symptoms and PCR-testing within 10 days of symptom onset, to account for reduced sensitivity of PCR-testing beyond this period. Individuals who had previously tested positive (PCR or antibody) prior to vaccination were excluded from the analysis.

Prior to May 2021, the Alpha variant was the main COVID-19 variant circulating in the UK after which time the Delta variant predominated. Cases were assigned to Alpha or Delta variant first on whole-genome sequencing, second on the S-gene target status (Alpha - negative prior to 28 June 2021; Delta - positive from 12 April 2021), third, for cases where sequencing or S-gene testing was not done, based on time-period: Alpha from 04 January 2021 to 02 May 2021 and Delta from 24 May 2021 as these variants were responsible for >80% of cases in all weeks during this period (>95% in most weeks).^11^

Testing data were linked to the Emergency Care Dataset (ECDS) to assess vaccine effectiveness against hospitalisation. We included emergency care attendances among symptomatic cases within 14 days of the positive test, which were not injury related, and resulted in an inpatient admission. ECDS data include hospital admissions through NHS emergency departments in England but not elective admissions. Only first attendances in the 14-day period were selected if a person had more than one admission from Emergency Care. Data were extracted on 08 September 2021 with cases included if tested by 03 September 2021. Sensitivity analysis was conducted using only admissions with COVID-19 or respiratory SNOMED CT codes as described in Public Health England’s Emergency department: weekly bulletin.^12^

When assessing effectiveness against death, data from the National Immunisation Management System on deaths provided by NHS Digital was used.

### Statistical analysis

We have previously reported the methodology for test-negative case-control analysis.^*1*^ Vaccine effectiveness was adjusted in logistic regression models for age, sex, index of multiple deprivation, ethnic group, care home residence status (for analyses including adults aged >=65 years), geographic region, period (calendar week), health and social care worker status (for analyses with adults aged <65 years), and clinical risk group (only available for <65 year-olds) or a clinically extremely vulnerable group (any age).^13^ For deaths, the period was modelled using a cubic spline due to smaller numbers.

Analyses were stratified by age-group according to the timing of vaccination in the general population as described in Table S1. Within age 65 years and above the analyses were also further stratified for those flagged as being in a clinical clinically extremely vulnerable and within age 40-64 for those flagged as being either clinically extremely vulnerable or in a risk group.

Vaccine effectiveness was assessed for each vaccine separately and by intervals after vaccination of at least 28 days post first dose, and at least 14 days post second dose. To assess potential waning, intervals of 1 week (7-13 days), 2-9 weeks, 10-14 weeks, 15-19 weeks and over 20 weeks were used. An additional analysis was done in the 80 years an older group according to interval between doses (≤28 days, ≥56 days) for the hospitalisation outcome and with an additional over 25 weeks post dose 2 interval. Where sample size was very small and VE estimates highly imprecise, estimates are not presented. This mainly affected the 1-week interval after the second dose and for longer intervals after the second dose effectiveness against Alpha and in younger ages effectiveness against Delta. Waning was not assed for age 16-39 as there was insufficient follow-up time in this group.

## Results

### Descriptive Statistics and characteristics

A total of 1,475,391 symptomatic individuals with symptom onset recorded within 10 days of a sample date had a first recorded positive SARS-CoV-2 test result during the study period, of whom 543,630 were classified as Alpha, 894,965 Delta and 36,796 as other/unknown (not included in the effectiveness analysis). A total of 3,757,981 negative tests were included from 3,299,344 individuals (402,136 were second negative results in the same individuals and 56,501 third negatives results, all >7 days after a previous negative). A description of the test positive and negative cases is given in Supplementary Tables S2 and S3. Overall, 2,025,292 (38.7%) had two doses of Vaxzevria, 1,659,513 (31.7%) had two doses of Comirnaty, 124,934 (2.4%) had two doses of Spikevax and 9,563 (0.2%) had a mixed course of two different vaccines or an interval of <19 days between first and second doses (Supplementary Table S2). Individuals with a mixed course and very short interval were excluded from analyses. Of the positive cases included in the analysis, 20,754 (1.4%) were hospitalised within 14 days of the test and 4,540 (0.3%) died within 28 days (Supplementary Table S3).

### Vaccine effectiveness estimates and vaccine waning

Vaccine effectiveness by dose, vaccine and age-group for the different outcomes is summarised in Supplementary Table S4. In general, higher vaccine effectiveness is seen with the mRNA vaccines compared to Vaxzevria, against the more severe outcomes compared to symptomatic infection, with Alpha compared to Delta and in younger compared to older age groups.

Table 1 and Figure 1 summarise vaccine effectiveness against symptomatic disease by week after the second dose for the Delta variant. Vaccine effectiveness against symptomatic disease peaked in the early weeks after the second dose then fell to 47.3 (95% CI 45.0 to 49.6) and 69.7 (95% CI 68.7 to 70.5)) by 20+ weeks against the Delta variant for Vaxzevria and Comirnaty, respectively. Waning of vaccine effectiveness was greater among 65+ year-olds compared to 40-64 year-olds. There was insufficient follow-up to estimate waning among <40 year-olds who were vaccinated more recently. Follow-up data after infection with the Alpha variant was also limited because this variant stopped circulating by the time the later follow-up periods were reached (Supplementary Tables S4 to S6).

**Table 1.** Vaccine effectiveness against Delta symptomatic disease among individuals with two doses of Vaxzevria, Comirnaty or Spikevax in England at 1 week, 2-9 weeks, 10 -14 weeks, 15-19 weeks and 20+ weeks.

|  |  | Dose 2 |  |  |  |  |
| --- | --- | --- | --- | --- | --- | --- |
| Vaccine | Age group | week 1 | 2-9 weeks | 10-14 weeks | 15-19 weeks | 20+ weeks |
| Vaxzevria | 16+ | 62.7 (61.7 to 63.8) | 66.7 (66.3 to 67.0) | 59.3 (58.8 to 59.9) | 52.6 (51.7 to 53.5) | 47.3 (45.0 to 49.6) |
|  | 65+ | 63.8 (48.2 to 74.8) | 58.9 (54.8 to 62.6) | 49.9 (45.4 to 54.0) | 43.3 (38.1 to 48.0) | 36.6 (28.7 to 43.7) |
|  | 40-64 | 57.1 (55.5 to 58.6) | 63.6 (62.9 to 64.3) | 59.8 (58.8 to 60.7) | 56.9 (55.3 to 58.4) | 57.8 (50.9 to 63.7) |
|  | 16-39 | 62.2 (52.5 to 70.0) | 65.5 (60.9 to 69.5) |  |  |  |
| Comirnaty | 16+ | 92.4 (92.1 to 92.7) | 89.8 (89.6 to 90.0) | 80.3 (79.9 to 80.6) | 73.4 (72.9 to 73.9) | 69.7 (68.7 to 70.5) |
|  | 65+ | 65.4 (34.2 to 81.8) | 80.1 (77.5 to 82.4) | 69.1 (66.2 to 71.8) | 62.1 (58.6 to 65.4) | 55.3 (50.2 to 60.0) |
|  | 40-64 | 87.9 (86.1 to 89.4) | 84.9 (84.3 to 85.4) | 78.2 (77.5 to 78.9) | 74.2 (73.1 to 75.3) | 75.7 (71.1 to 79.5) |
|  | 16-39 | 92.5 (92.1 to 92.8) | 91.0 (90.8 to 91.3) | 77.1 (71.4 to 81.6) |  |  |
| Spikevax | 16+ | 95.2 (94.4 to 95.9) | 94.5 (94.1 to 95.0) | 90.3 (67.2 to 97.1) |  |  |
|  | 40-64 | 94.0 (92.1 to 95.5) | 93.7 (92.9 to 94.4) | 96.1 (70.1 to 99.5) |  |  |
|  | 16-39 | 95.0 (94.1 to 95.8) | 94.9 (94.2 to 95.5) |  |  |  |

**Figure 1.**
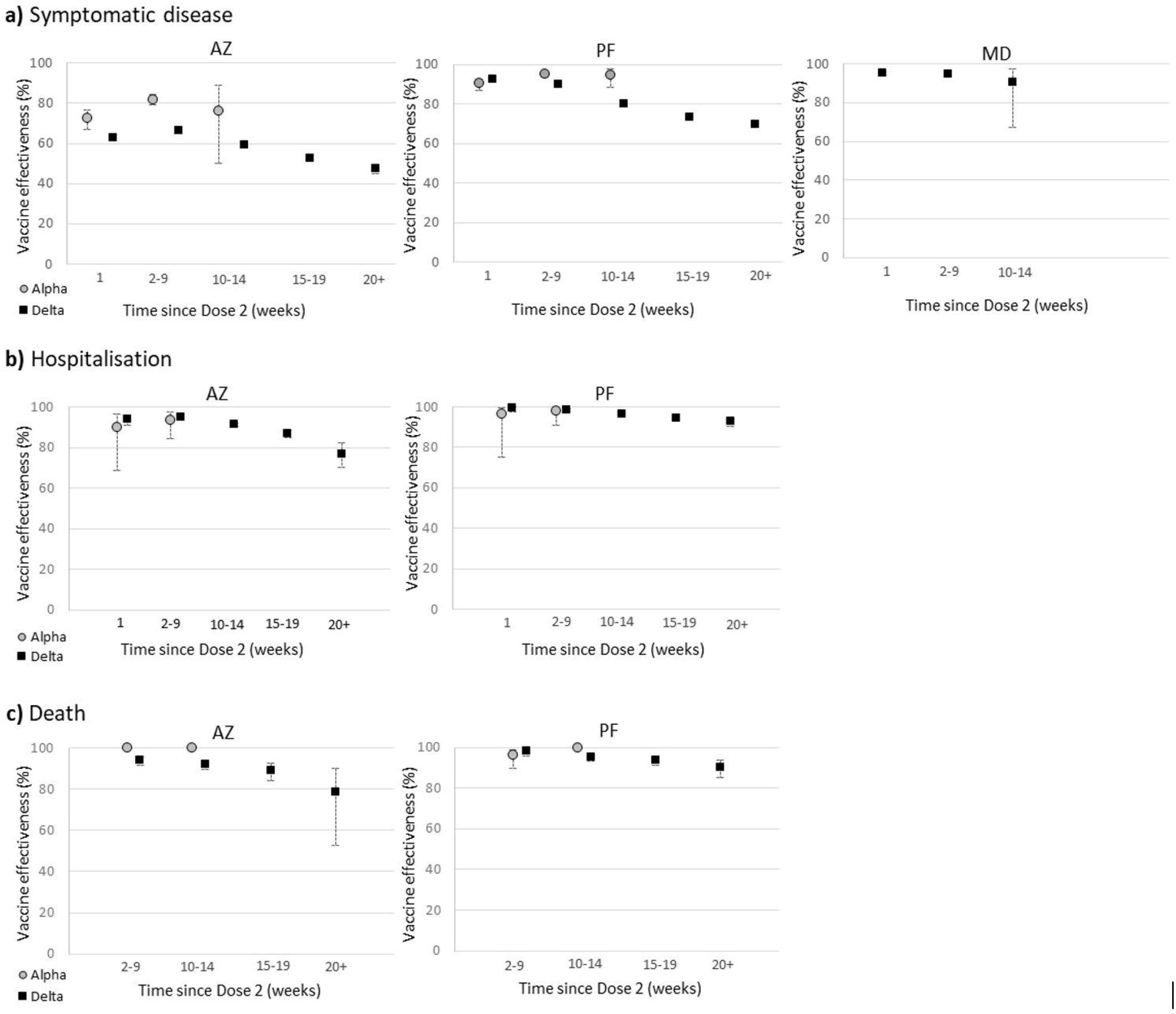
Vaccine effectiveness against Delta symptomatic disease, hospitalisation and death among individuals aged 16+ with two doses of Vaxzevria (AZ), Comirnaty (PF) or Spikevax (MD) in England and 95% confidence intervals.

There was limited waning in protection against hospitalization, with a vaccine effectiveness of 77.0 (70.3 to 82.3) and 92.7 (90.3 to 94.6) beyond 20 weeks post-vaccination for Vaxzevria and Comirnaty, respectively (Delta only) (Figure 1 and Table 2). Combined results for both vaccines are shown in Supplementary Figure S3. Vaccine effectiveness against hospitalisation for the Alpha variant is reported in Supplementary Table S6. Similarly, there was limited waning of vaccine effectiveness against deaths Vaxzevria (VE 78.7 (95% CI 52.7 to 90.4)) and Comirnaty (VE 90.4 (95% CI 85.1 to 93.8)) beyond 20 weeks post-vaccination for all ages (Figure 1 and Table 3).

**Table 2.**
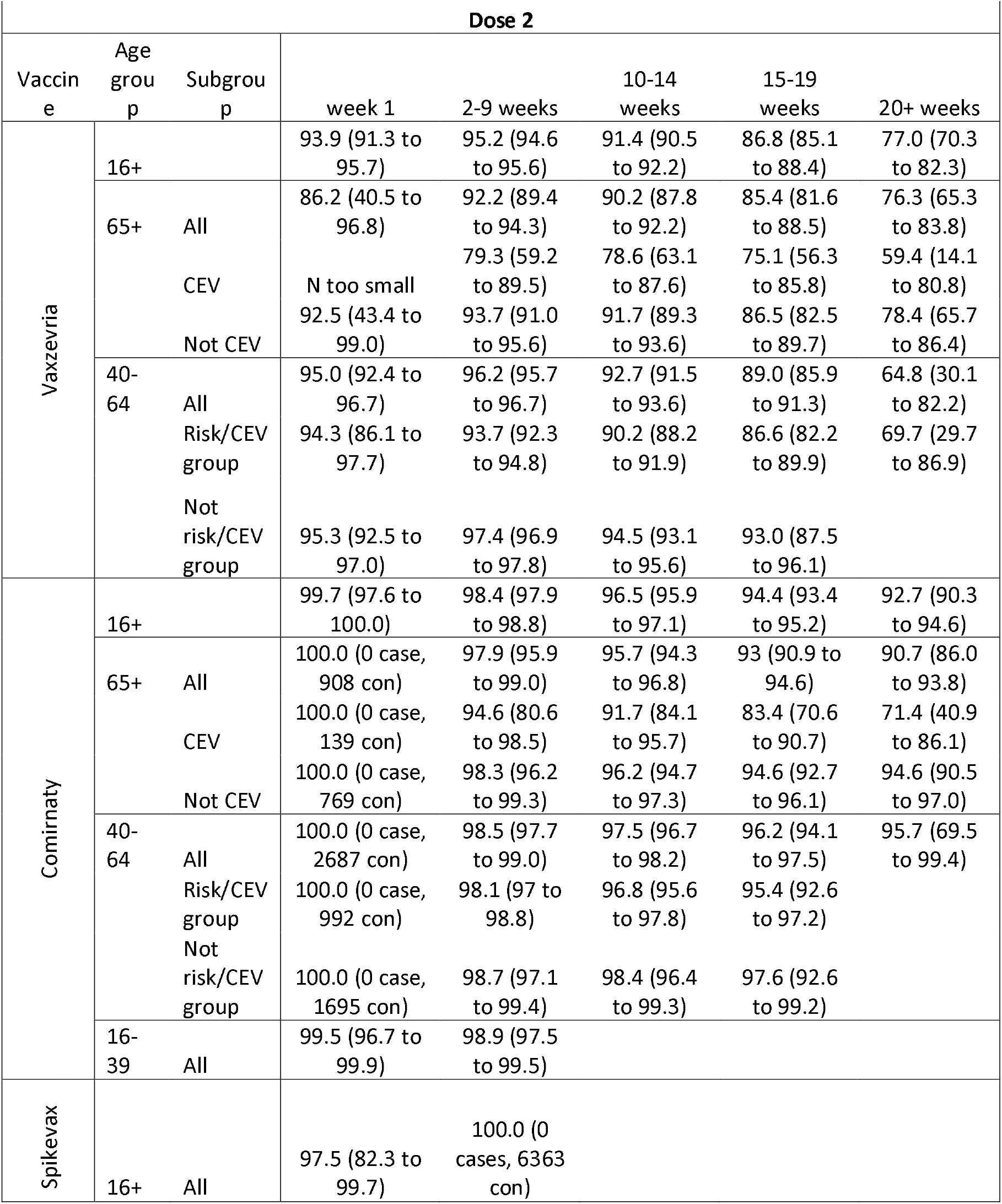
Vaccine effectiveness against Delta hospitalisation among individuals with two doses of Vaxzevria, Comirnaty or Spikevax in England at 1 week, 2-9 weeks, 10 -14 weeks, 15-19 weeks and 20+ weeks.

**Table 3.**
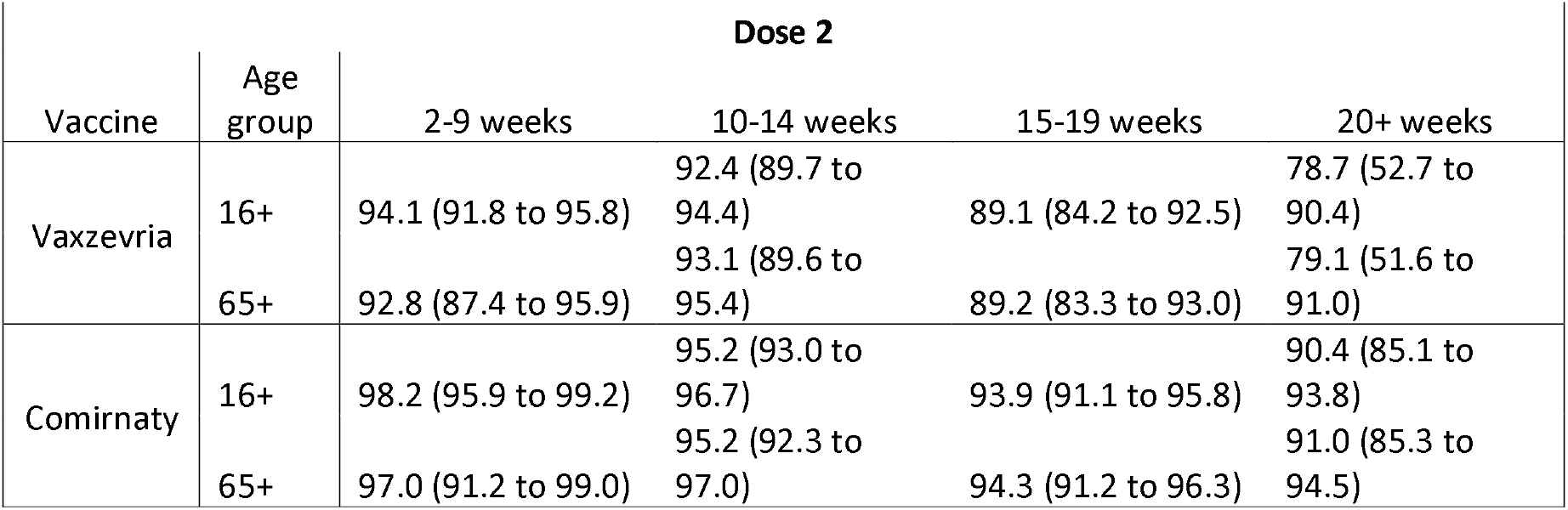
Vaccine effectiveness against Delta deaths among individuals with two doses of Vaxzevria, Comirnaty in England at 2-9 weeks, 10 -14 weeks, 15-19 weeks and 20+ weeks.

In general, there was greater waning in protection against hospitalisation among the oldest age-group, except in the 20+ weeks period with Vaxzevria, though there was limited data in the 40-64 years group for this period and confidence intervals overlapped (Table2, Figure 2). Results from the sensitivity analysis using only admissions coded as respiratory admissions showed similar effectiveness against hospitalisation compared to all hospitalisation codes, with vaccine effectiveness estimates of 97.4 (95% CI 97 to 97.7) for Comirnaty and 93.9 (95% CI 93.4 to 94.4) for Vaxzevria for the Delta variant in the 20+ weeks period.

**Figure 2.**
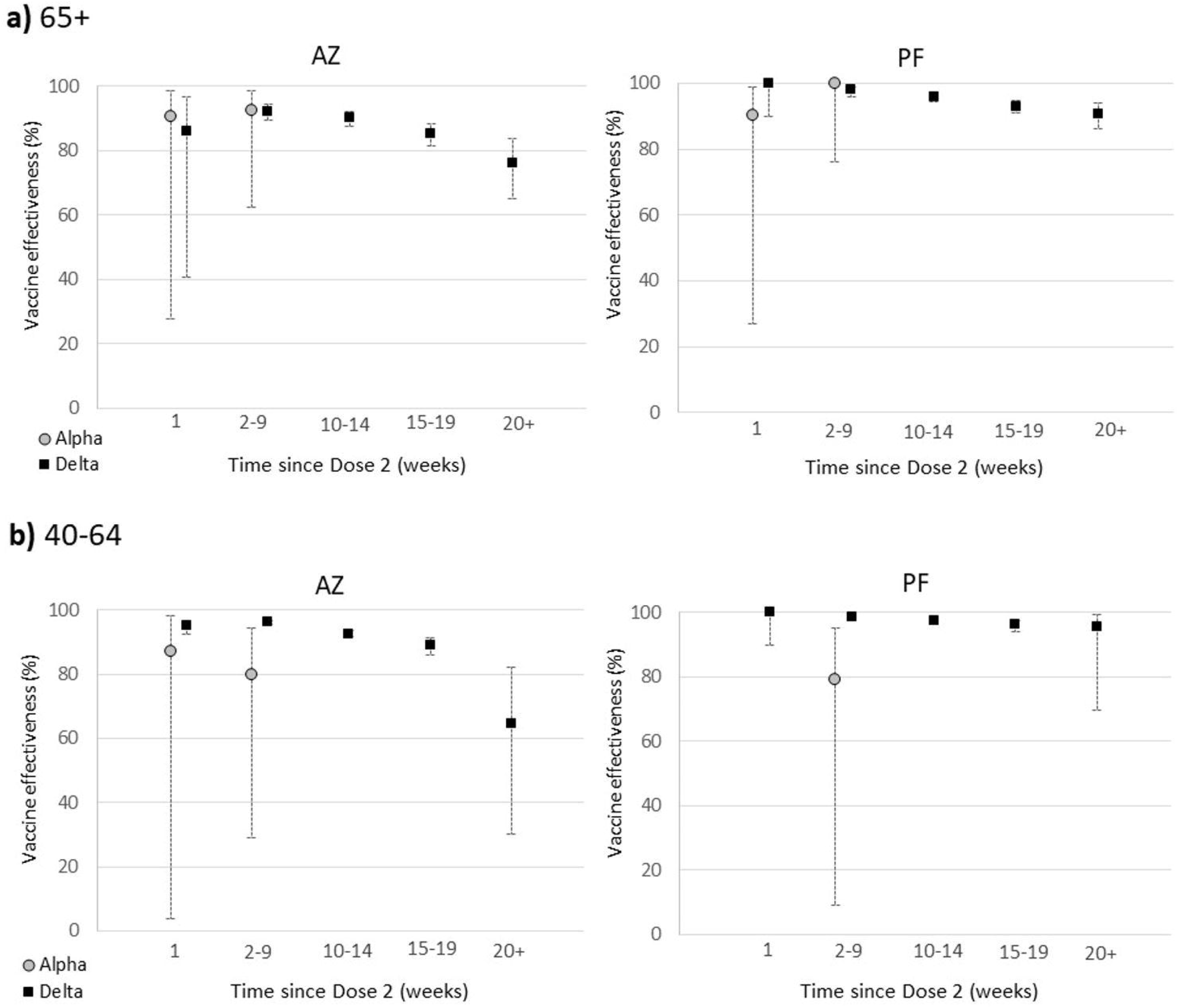
Vaccine effectiveness against hospitalisation by age group for Vaxzevria (AZ) and Comirnaty (PF), for a) 65+ years and b) 40-64 years.

Stratifying by risk group status identified greater waning among 65+ year-olds in a clinically extremely vulnerable group and among 40-64-year olds in a clinical risk group or clinically extremely vulnerable group, compared to those who were not in the group (Figure 3 and Supplementary Figure S1). For those aged 40-64 years, recipients of either vaccine who were not in a risk group had very high vaccine effectiveness against hospitalisation throughout the follow up period (Figure S1). There was also very little evidence of waning up to 20+ weeks post-vaccination among 65+ year-olds who were not in a CEV group and had received Comirnaty (Figure 3).

**Figure 3.**
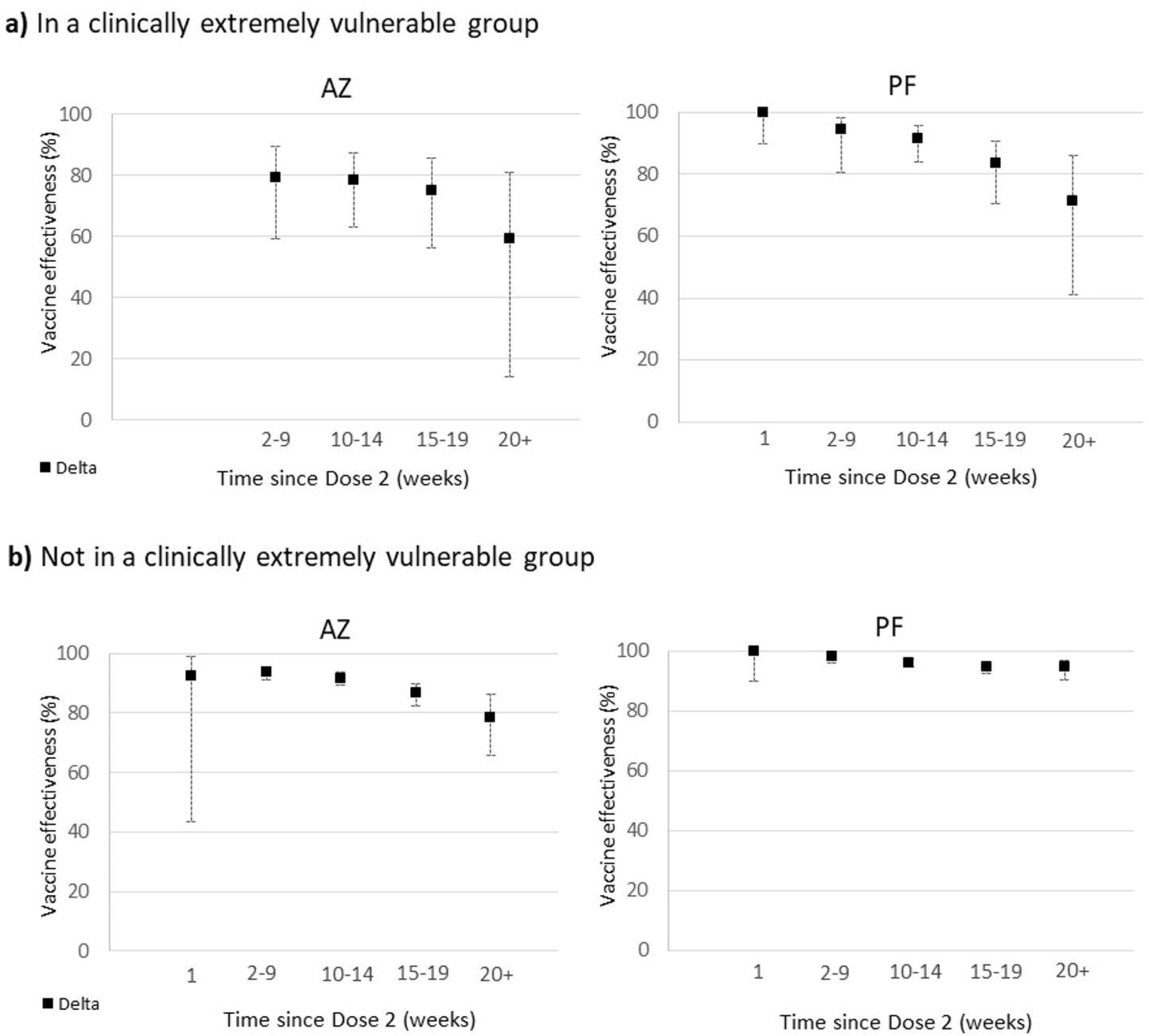
Vaccine effectiveness against hospitalisation (age >=65 years) by clinical extremely vulnerable group status for Vaxzevria (AZ) and Comirnaty (PF).

An analysis restricted to 80+ year-olds who had received Comirnaty prior to 04 January 2021 found lower vaccine effectiveness among those with a short (<4 week), compared to an extended interval (≥ 8 weeks) between doses in the latest follow-up periods (beyond 20 weeks after dose 2), although confidence intervals were wide and overlapping (Supplementary Figure S2).

## DISCUSSION

### Key findings

Our data provide evidence of waning of protection against symptomatic infection following both Vaxzevria and Comirnaty vaccines from 10 weeks after the second dose. Protection against hospitalisation and death, however, was sustained at very high levels for at least 20 weeks after the second dose. Beyond 20 weeks, we observed more waning with Vaxzevria compared to Comirnaty, though the groups who received either vaccine differed. Waning of protection against hospitalisation was greater among older adults and in those in a clinical risk group. Among ≥65 year-olds who were not in a clinical risk group, however, protection against hospitalisation remains close to 95% with Comirnaty and just under 80% with Vaxzevria beyond 20 weeks after the second vaccine dose.

### Comparison with existing literature

Our finding of waning in vaccine effectiveness against symptomatic disease is consistent with recent findings from Israel and Qatar reporting an increasing proportion of breakthrough cases among the earliest vaccinated individuals.^9,14-16^ In addition to the emergence and rapid spread of the more transmissible Delta variant, the observed waning protection against symptomatic infection with time since vaccination may also be contributing to the increase in COVID-19 cases seen in the UK and elsewhere. Reassuringly, though the number of hospitalisations and, particularly, deaths due to COVID-19 have remained low, especially among vaccinated adults.^17^ Our finding of only limited waning of protection against hospitalisation or death in most of the groups studied is consistent with the preserved vaccine effectiveness against hospitalisation reported in Qatar.^9^ Studies from the US have also reported sustained high VE against hospitalisation due to COVID-19 despite the emergence and rapid spread of the Delta variant in the community. Across 18 US states, VE following two doses given 3 weeks apart among adults aged ≥18 years (median age 59 years) admitted to 21 hospitals during 11 March to 14 July 2021, was 86% (95% CI = 82%–88%) overall, and 87% (95% CI, 83%–90%) in patients with illness onset during March–May compared to 84% (95% CI, 79%–89%) in those with illness onset during June–July 2021, with no evidence of a significant decline in VE over the 24-week period.^18^ A similar study of New York adults during 3 May to 25 July 2021 found hospitalisation rates to be nearly 10-fold lower in vaccinated (>90% received 2 mRNA doses 3 weeks apart) compared to unvaccinated adults (1.31 vs. 10.69 per 100,000 person-days). VE against hospitalisation remained relatively stable (91.9%–95.3%) during the surveillance period, although age-adjusted VE against new COVID-19 cases declined from 91.7% to 79.8%, coinciding with an increase in Delta variant circulation from <2% to >80%.^19^ Conversely, there have been reports of an increased proportion of hospitalised cases among the earliest vaccinated adults receiving two doses of Cominarty three weeks apart in Israel.^14^ The shorter dosing interval of 3 weeks and, hence, longer follow-up of a population with rapid vaccine uptake in Israel may be a factor in explaining this difference compared to the UK.

### Implications

Our findings and those of Qatar and the US raise important questions about the timing of additional boosters in fully vaccinated adults who remain protected against hospitalisation and death for at least 5 months after vaccination. Israel was one of the first countries to immunise adults with the Cominarty and began offering a third dose of the same vaccine to older adults from the end of July 2021.^20^ Early data indicate that the third dose was associated with large reductions in SARS-CoV-2 infection within one week of vaccination, with greater reductions in the second week.^20^ The duration of protection offered by the third dose, however, remains to be established. The US, the UK and other countries are currently considering recommendations for booster doses for specific age-groups and risk-groups.

Booster doses improve both humoral and cellular immunity against SARS-CoV-2 and have increased neutralising activity against the different variants including Delta, which is likely to improve protection against infection. Given the sustained high VE against hospitalisation and death, the additional benefit of a third dose against these more serious outcomes is limited in the current epidemiological situation. VE may, however, continue to wane over time and it is likely that a booster doses may have a bigger impact on the more severe outcomes with longer intervals between the second and third doses. Decisions on the timing of the third dose will have to balance the rate of waning immunity against disease epidemiology, including the emergence of variants, and prioritisation of those at highest risk of waning immunity. At the same time, it is likely that the third dose will be more reactogenic than previous doses, especially if primed and boosted with different vaccines.^21^ Attractive alternatives include half-dose boosters or boosting with variant SARS-CoV-2 strains, which are both under investigation currently.^22^

For the UK and countries such as Canada, another important consideration is that the extended interval schedules of 8-12 weeks provide higher serological responses and increased vaccine effectiveness compared to the licensed 3-week interval for mRNA vaccines,^23^ thus potentially providing their populations with better longer-term protection.^24^ This is supported by our findings comparing short and long intervals in 80+ year olds in the current analysis.

We found that waning was greatest among individuals in clinical risk groups, suggesting that this group should be prioritised for boosters, whenever they are recommended. Other studies have reported lower immune responses and vaccine effectiveness among individuals in clinical risk groups, most notably among immunosuppressed groups.^10,18,25,26^ The UK and others have recently recommended a third dose of COVID-19 vaccine for immunocompromised adults as part of their primary immunisation course because of lower immunogenicity following two doses, with improved responses after 3 doses. ^27,28^

### Limitations

The strengths and limitations of our dataset and an evaluation of vaccine effectiveness using the test negative case control design have been described in detail previously.^1,6^ There are some notable limitations worth highlighting that could particularly affect the interpretation of results on waning. First, there are important differences in the groups that received different vaccines as part of the roll-out of the national programme in the UK. For example, Comirnaty was less likely to be used in care homes and among individuals in clinical risk groups, in particular housebound individuals, due to challenges in ultra-low temperature storage and delivery. Instead, Vaxzevria was more likely to be given to these groups but this vaccine was not given to healthy adults under 40 years of age following a recommendation to use alternative vaccines in this age-group because of the risk of vaccine-induced thrombotic thrombocytopenia.^29^ While we do adjust for age-group and clinical risk groups in our analysis, this is unlikely to fully mitigate the differences. Second, over time there will be an increasing number of individuals who would have been infected both prior to and after vaccination. Where they have previously tested positive they will be excluded from the analysis, but many will remain unknown. This means that an increasing proportion of the unvaccinated control group may have some level of protection from natural infection. This will attenuate vaccine effectiveness over time. Third, testing policies have changed over time. For example, PCR is increasingly being used to confirm positive self-administered lateral flow tests. There is an increased risk of false negative PCRs among those that were lateral flow positive, which will result in misclassification in our study based on PCR results. This would again attenuate vaccine effectiveness.

## Conclusions

Our study provides evidence of significant waning against symptomatic disease but limited waning against severe disease over a period of at least 5 after administration of second doses in a programme with an extended interval between first and second doses. Waning appeared to be greater in older age groups and among individuals in clinical risk groups, suggesting that these individuals should be prioritised for booster doses.

## Data Availability

Data may be available upon request.

## References

1. Lopez Bernal J, Andrews N, Gower C, et al. Effectiveness of Covid-19 Vaccines against the B.1.617.2 (Delta) Variant. The New England Journal of Medicine 2021;385:585–94.

2. Sharif A. Ismail TGV, Suzanne Elgohari, Julia Stowe, Elise Tessier, Nick Andrews, Amoolya Vusirikala, Mary Ramsay, Sema Mandal, Jamie Lopez Bernal. Effectiveness of BNT162b2 mRNA and ChAdOx1 adenovirus vector COVID-19 vaccines on risk of hospitalisation among older adults in England: an observational study using surveillance data. 2021.

3. Vasileiou E, Simpson CR, Shi T, et al. Interim findings from first-dose mass COVID-19 vaccination roll-out and COVID-19 hospital admissions in Scotland: a national prospective cohort study. The Lancet 2021;397:1646–57.

4. Pritchard E, Matthews PC, Stoesser N, et al. Impact of vaccination on SARS-CoV-2 cases in the community: a population-based study using the UK’s COVID-19 Infection Survey. medRxiv 2021:2021.04.22.21255913.

5. Hyams C, Marlow R, Maseko Z, et al. Effectiveness of BNT162b2 and ChAdOx1 nCoV-19 COVID-19 vaccination at preventing hospitalisations in people aged at least 80 years: a test-negative, case-control study. The Lancet Infectious Diseases.

6. Lopez Bernal J, Andrews N, Gower C, et al. Effectiveness of the Pfizer-BioNTech and Oxford-AstraZeneca vaccines on covid-19 related symptoms, hospital admissions, and mortality in older adults in England: test negative case-control study. BMJ 2021;373:n1088.

7. Pouwels KB, Pritchard E, Matthews PC, et al. Impact of Delta on viral burden and vaccine effectiveness against new SARS-CoV-2 infections in the UK. medRxiv 2021:2021.08.18.21262237.

8. Nanduri S PT, Derado G, et al. Effectiveness of Pfizer-BioNTech and Moderna Vaccines in Preventing SARS-CoV-2 Infection Among Nursing Home Residents Before and During Widespread Circulation of the SARS-CoV-2 B.1.617.2 (Delta) Variant — National Healthcare Safety Network, March 1–August 1, 2021. MMWR Morb Mortal Wkly Rep 2021;70:1163–6.

9. Chemaitelly H, Tang P, Hasan MR, et al. Waning of BNT162b2 vaccine protection against SARS-CoV-2 infection in Qatar. medRxiv 2021:2021.08.25.21262584.

10. Shrotri M, Fragaszy E, Geismar C, et al. Spike-antibody responses to ChAdOx1 and BNT162b2 vaccines by demographic and clinical factors (Virus Watch study). medRxiv 2021:2021.05.12.21257102.

11. Public Health England. SARS-CoV-2 variants of concern and variants under investigation in England Technical briefing 22. 2021.

12. Emergency department: weekly bulletins for 2021. 2021. (Accessed 13/09/2021, at https://www.gov.uk/government/publications/emergency-department-weekly-bulletins-for-2021.)

13. COVID-19 — high risk shielded patient list identification methodology: rule logic. 2020. 13/09/2021, at https://digital.nhs.uk/coronavirus/shielded-patient-list/methodology/rule-logic.)

14. Goldberg Y, Mandel M, Bar-On YM, et al. Waning immunity of the BNT162b2 vaccine: A nationwide study from Israel. medRxiv 2021:2021.08.24.21262423.

15. Israel A, Merzon E, Schäffer AA, et al. Elapsed time since BNT162b2 vaccine and risk of SARS-CoV-2 infection in a large cohort. medRxiv 2021:2021.08.03.21261496.

16. Mizrahi B, Lotan R, Kalkstein N, et al. Correlation of SARS-CoV-2 Breakthrough Infections to Time-from-vaccine; Preliminary Study. medRxiv 2021:2021.07.29.21261317.

17. Public Health England. Weekly national Influenza and COVID-19 surveillance report Week 36 report (up to week 35 data) 9 September 2021. 2021.

18. Mark W. Tenforde M, PhD1,*; Wesley H. Self, MD2,*; Eric A. Naioti, MPH1; Adit A. Ginde, MD3; David J. Douin, MD3; Samantha M. Olson, MPH1;, H. Keipp Talbot MJDC, MD2; Nicholas M. Mohr, MD4; Anne Zepeski, PharmD4; Manjusha Gaglani, MBBS5,6; Tresa McNeal, MD5;, Shekhar Ghamande MNIS, MD7; Kevin W. Gibbs, MD8; D. Clark Files, MD8; David N. Hager, MD, PhD9; Arber Shehu, MD9;, et al. Sustained Effectiveness of Pfizer-BioNTech and Moderna Vaccines Against COVID-19 Associated Hospitalizations Among Adults — United States, March–July 2021. Morbidity and Mortality Weekly Report (MMWR) 2021;70.

19. Eli S. Rosenberg DRH, Vajeera Dorabawila, MaryBeth Conroy, Danielle Greene, Emily Lutterloh, Bryon Backenson, Dina Hoefer, Johanne Morne, Ursula Bauer, Howard A. Zucker. New COVID-19 Cases and Hospitalizations Among Adults, by Vaccination Status — New York, May 3–July 25, 2021. Morbidity and Mortality Weekly Report (MMWR) 2021;70.

20. Israel’s COVID-19 boosters are preventing infections, new studies suggest. 2021. (Accessed 13/09/2021, at https://www.science.org/content/article/israel-s-covid-19-boosters-are-preventing-infections-new-studies-suggest.)

21. Powell AA, Power L, Westrop S, et al. Real-world data shows increased reactogenicity in adults after heterologous compared to homologous prime-boost COVID-19 vaccination, March−June 2021, England. 2021;26:2100634.

22. Wu K, Choi A, Koch M, et al. Preliminary Analysis of Safety and Immunogenicity of a SARS-CoV-2 Variant Vaccine Booster. medRxiv 2021:2021.05.05.21256716.

23. Amirthalingam G, Bernal JL, Andrews NJ, et al. Higher serological responses and increased vaccine effectiveness demonstrate the value of extended vaccine schedules in combatting COVID-19 in England. medRxiv 2021:2021.07.26.21261140.

24. Khoury DS, Cromer D, Reynaldi A, et al. Neutralizing antibody levels are highly predictive of immune protection from symptomatic SARS-CoV-2 infection. Nature medicine 2021;27:1205–11.

25. Heather J Whitaker RST, Rachel Byford, Nick J Andrews, Julian Sherlock, Praveen Sebastian Pillai, John Williams, Elizabeth Button, Helen Campbell, Mary Sinnathamby, Georgina Pike, Sneha Anand, Ezra Linley, Jacqueline Hewson, Ashley D Otter, Joanna Ellis, Richard FD Hobbs, Maria Zambon5, Mary Ramsay, Kevin E Brown, Simon de Lusignan, Gayatri Amirthalingam, Jamie Lopez Bernal. Pfizer-BioNTech and Oxford AstraZeneca COVID-19 vaccine effectiveness and immune response among individuals in clinical risk groups. 2021.

26. Monin L, Laing AG, Muñoz-Ruiz M, et al. Safety and immunogenicity of one versus two doses of the COVID-19 vaccine BNT162b2 for patients with cancer: interim analysis of a prospective observational study. The Lancet Oncology 2021;22:765–78.

27. Werbel WA, Boyarsky BJ, Ou MT, et al. Safety and Immunogenicity of a Third Dose of SARS-CoV-2 Vaccine in Solid Organ Transplant Recipients: A Case Series. Annals of Internal Medicine 2021.

28. Longlune N, Nogier MB, Miedougé M, et al. High immunogenicity of a messenger RNA based vaccine against SARS-CoV-2 in chronic dialysis patients. Nephrology, dialysis, transplantation : official publication of the European Dialysis and Transplant Association - European Renal Association 2021.

29. Public Health England. COVID-19: the green book, chapter 14a Coronavirus (COVID-19) vaccination information for public health professionals. 2021.

