## Supplementary Table S1 for "Vaccine effectiveness and duration of protection of Comirnaty, Vaxzevria and Spikevax against mild and severe COVID-19 in the UK"

**Table S1:** Analysis sets by age and outcomes

| Age at March 31st 2021 | Outcome | Period for cases / controls included  (Alpha)* | Period for cases/controls included  (Delta)** | Period in which first vaccination dose is given if vaccinated at onset |
| --- | --- | --- | --- | --- |
| 80+ | Onset of Symptomatic infection | Onset  Dec 8 2020 – June 27 2021 | Onset from Apr 12 2021 and tested by Sep 3 2021 | Dec 8 2020 – Jan 3 2021 |
|  | Hospitalisation within 14 days of test | Onset  Dec 8 2020 – June 27 2021 | Onset from Apr 12 2021 and tested by  Sep 3 2021 | Dec 8 2020 – Jan 3 2021 |
| 65+ | Onset Symptomatic infection | Onset Jan 4 2021 – June 27 2021 | Onset from Apr 12 2021 and tested by Sep 3 2021 | Jan 4 2021 onwards |
|  | Hospitalisation  within 14 days of test | Onset Jan 4 2020 – June 27 2021 | Onset from Apr 12 2021 and tested by Sep 3 2021 | Jan 4 2021 onwards |
|  | Death within 28 days of test | Onset Jan 4 2021 – June 27 2021 | Onset from Apr 12 2021 and tested by Sep 3 2021 | Jan 4 2021 onwards |
| 40-64 | Onset of Symptomatic infection | Onset Feb 1 2020-June 27 2021 | Onset from Apr 12 2021 and tested by Sep 3 2021 | Feb 1 2021 onwards |
|  | Hospitalisation within 14 days of test | Onset Feb 1 2020-June 27 2021 | Onset from Apr 12 2021 and tested by Sep 3 2021 | Feb 1 2021 onwards |
| 16-39 | Onset of Symptomatic infection | Onset May 10 2020-June 27 2021 | Onset from May 10 2021 and tested by Sep 3 2021 | May 10 2021 onwards |
|  | Hospitalisation within 14 days of test | Onset May 10 2020-June 27 2021 | Onset from May 10 2021 and tested Sep 3 2021 | May 10 2021 onwards |
| 16+ (All) | Onset of Symptomatic infection | Onset Jan 4 2021 – June 27 2021 | Onset from Apr 12 2021 and tested by Sep 3 2021 | Jan 4 2021 onwards |
|  | Hospitalisation within 14 days of test | Onset Jan 4 2020 – June 27 2021 | Onset from Apr 12 2021 and tested by Sep 3 2021 | Jan 4 2021 onwards |
|  | Death within 28 days of test | Onset Jan 4 2021 – June 27 2021 or for 80+ cohort Dec 8 2021-June 27 2021 | Onset from Apr 12 2021 and tested by Sep 3 2021 | Jan 4 2021 onwards if aged <80 or Dec 8 2021-Jan 3 2021 if aged 80+ |

*June 27 is for test negative controls and test positive cases if sequencing or s-gene target failure is done, otherwise May 2 is used before which >80% of those tested positive were Alpha

**Apr 12 is for test negative controls and test positive cases if sequencing or s-gene target failure is done , otherwise May 24 is used after which >80% of those tested positive were Delta
