## Supplementary Table S2 for "Vaccine effectiveness and duration of protection of Comirnaty, Vaxzevria and Spikevax against mild and severe COVID-19 in the UK"

**Table S2.** Descriptive characteristics of positive and negative test results in individuals tested for SARS-CoV-2 in England for the study population. *

|  |  | **Overall** | | **Positive** | | **Negative** | |
| --- | --- | --- | --- | --- | --- | --- | --- |
|  |  | **n** | **%** | **n** | **%** | **n** | **%** |
|  | **Test Result** | **5,233,372** | **100.0** | **1,475,391** | **28.2%** | **3,757,981** | **71.8%** |
| **Vaccination status** | Unvaccinated | 806,829 | 15.4% | 337,142 | 22.9% | 469,687 | 12.5% |
|  | Vaxzevria 1 dose | 75,466 | 1.4% | 23,742 | 1.6% | 51,724 | 1.4% |
|  | Vaxzevria 2 doses | 2,025,292 | 38.7% | 525,721 | 35.6% | 1,499,571 | 39.9% |
|  | Comirnaty 1 dose | 474,266 | 9.1% | 191,606 | 13.0% | 282,660 | 7.5% |
|  | Comirnaty 2 doses | 1,659,513 | 31.7% | 349,171 | 23.7% | 1,310,342 | 34.9% |
|  | Spikevax 1 dose | 57,509 | 1.1% | 21,535 | 1.5% | 35,974 | 1.0% |
|  | Spikevax 2 doses | 124,934 | 2.4% | 24,328 | 1.6% | 100,606 | 2.7% |
|  | Mixed course or dose1-2 interval <19 days | 9,563 | 0.2% | 2,146 | 0.1% | 7,417 | 0.2% |
|  | 80+ | 66,725 | 1.3% | 19,517 | 1.3% | 47,208 | 1.3% |
| **Age Group** | 65-79 | 284,030 | 5.4% | 68,189 | 4.6% | 215,841 | 5.7% |
|  | 40-64 | 1,942,666 | 37.1% | 517,105 | 35.0% | 1,425,561 | 37.9% |
|  | 16-39 | 2,939,951 | 56.2% | 870,580 | 59.0% | 2,069,371 | 55.1% |
| **Gender** | female | 2,923,495 | 55.9% | 754,922 | 51.2% | 2,168,573 | 57.7% |
|  | male | 2,304,400 | 44.0% | 718,775 | 48.7% | 1,585,625 | 42.2% |
|  | missing | 5,477 | 0.1% | 1,694 | 0.1% | 3,783 | 0.1% |
| **Ethnicity** | African | 81,929 | 1.6% | 26,710 | 1.8% | 55,219 | 1.5% |
|  | Another Asian background | 67,789 | 1.3% | 21,289 | 1.4% | 46,500 | 1.2% |
|  | Another Black background | 9,431 | 0.2% | 3,170 | 0.2% | 6,261 | 0.2% |
|  | Another ethnic background | 39,912 | 0.8% | 11,198 | 0.8% | 28,714 | 0.8% |
|  | Arab | 22,505 | 0.4% | 6,418 | 0.4% | 16,087 | 0.4% |
|  | Bangladeshi | 46,185 | 0.9% | 17,431 | 1.2% | 28,754 | 0.8% |
|  | Caribbean | 43,118 | 0.8% | 16,490 | 1.1% | 266,284 | 7.1% |
|  | Chinese | 24,955 | 0.5% | 5,401 | 0.4% | 19,554 | 0.5% |
|  | Indian | 183,052 | 3.5% | 51,621 | 3.5% | 131,431 | 3.5% |
|  | Mixed or multiple ethnic groups | 112,294 | 2.1% | 34,119 | 2.3% | 78,175 | 2.1% |
|  | Pakistani | 139,249 | 2.7% | 46,741 | 3.2% | 92,508 | 2.5% |
|  | Prefer not to say | 176,641 | 3.4% | 50,412 | 3.4% | 126,229 | 3.4% |
|  | White | 4,286,312 | 81.9% | 1,184,391 | 80.3% | 3,101,921 | 82.5% |
| **NHS Region** | East of England | 572,833 | 10.9% | 146,863 | 10.0% | 425,970 | 11.3% |
|  | London | 769,829 | 14.7% | 209,676 | 14.2% | 560,153 | 14.9% |
|  | Midlands | 1,002,007 | 19.1% | 297,143 | 20.1% | 704,864 | 18.8% |
|  | North East | 871,790 | 16.7% | 274,149 | 18.6% | 597,641 | 15.9% |
|  | North West | 750,159 | 14.3% | 231,517 | 15.7% | 518,642 | 13.8% |
|  | South East | 770,922 | 14.7% | 191,248 | 13.0% | 579,674 | 15.4% |
|  | South West | 495,821 | 9.5% | 124,792 | 8.5% | 371,029 | 9.9% |
|  | Unknown | 11 | 0.0% | 3 | 0.0% | 8 | 0.0% |

*aged 80 and above and either unvaccinated at the time of onset or first vaccinated before January 4^th^, 2021 with onset December 8^th^, 2020-September 3rd, 2021 or aged 16 and above and unvaccinated prior to January 4^th^, 2021 with onset January 4^th^ 2021-September 3rd, 2021.
