## Supplementary Table S3 for "Vaccine effectiveness and duration of protection of Comirnaty, Vaxzevria and Spikevax against mild and severe COVID-19 in the UK"

**Table S3**. Counts by variant, hospitalisations and deaths for positive SARS-CoV-2 tests among individuals in England for the study population.

|  |  | **All** | **16-39** | **40-64** | **65-79** | **80+** |
| --- | --- | --- | --- | --- | --- | --- |
| **Variant** | **Alpha** | 543,630 | 260,209 | 241,854 | 31,795 | 9,772 |
|  | **Delta** | 894,965 | 594,370 | 261,019 | 34,175 | 5,401 |
|  | **Unknown** | 36,796 | 16001 | 14232 | 2219 | 4344 |
| **Outcome** | **Number of hospitalisations** | 20,754 | 4,766 | 9,856 | 3,757 | 2,375 |
|  | **Number of deaths** | 4,540 | 85 | 930 | 1203 | 2322 |
