## Supplementary Table S4 for "Vaccine effectiveness and duration of protection of Comirnaty, Vaxzevria and Spikevax against mild and severe COVID-19 in the UK"

**Table S4.** Vaccine effectiveness against Alpha and Delta symptomatic disease, hospitalisation and death for Comirnaty, Vaxzevria and Spikevax in England.

|  |  |  | Symptomatic disease (tested by Sep 3rd) | | Hospitalisation (tested by Sep 3rd) | | Death (tested by Sep 3rd) | |
| --- | --- | --- | --- | --- | --- | --- | --- | --- |
| Age | Vaccine | Dose* | Alpha | Delta | Alpha | Delta | Alpha | Delta |
| All | Comirnaty | 1 | 45.7 (44 to 47.3) | 51.9 (51.4 to 52.4) | 85.2 (81.6 to 88.1) | 91.8 (90.4 to 93) | 73.1 (65 to 79.3) | 88.6 (77.3 to 94.3) |
| from Jan 4 |  | 2 | 95.0 (93.8 to 95.9) | 83.5 (83.3 to 83.6) | 97.9 (91.4 to 99.5) | 96.7 (96.3 to 97) | 96.3 (89.9 to 98.6) | 95.2 (93.7 to 96.4) |
|  | Vaxzevria | 1 | 44.5 (42.9 to 46.1) | 43.3 (42.3 to 44.2) | 82.5 (78.7 to 85.7) | 81.4 (78.7 to 83.7) | 79.1 (68.8 to 86) | 88.4 (78.2 to 93.8) |
|  |  | 2 | 81.7 (79.0 to 84.0) | 65.2 (64.9 to 65.6) | 93.9 (84.9 to 97.5) | 93.0 (92.4 to 93.5) | 100 (0 case, 64,518 con) | 92.7 (90.7 to 94.3) |
|  | Spikevax | 1 | 54.5 (8.5 to 77.3) | 65.9 (65.0 to 66.7) |  | 95.2 (91.8 to 97.1) |  |  |
|  |  | 2 |  | 94.8 (94.4 to 95.2) |  |  |  |  |
|  | Any | 1 | 45.1 (43.9 to 46.3) | 51.8 (51.4 to 52.2) | 83.9 (81.2 to 86.2) | 88.4 (87.2 to 89.5) | 75.3 (68.7 to 80.5) | 88.3 (73.5 to 94.9) |
|  |  | 2 | 89.7 (88.4 to 90.8) | 73.3 (73.1 to 73.5) | 96.1 (91.5 to 98.2) | 94.4 (94.0 to 94.8) | 97.0 (91.7 to 98.9) | 93.0 (90.3 to 95.0) |
| 16-39 | Comirnaty | 1 |  | 52.6 (52.1 to 53.1) |  | 91.1 (89.4 to 92.5) |  |  |
| from May 10 |  | 2 |  | 91.1 (90.9 to 91.4) |  | 98.9 (97.6 to 99.5) |  |  |
|  | Vaxzevria | 1 |  | 48.5 (44.6 to 52.1) |  | 87.8 (67.4 to 95.4) |  |  |
|  |  | 2 |  | 66.0 (61.5 to 69.9) |  | 100.0 (no case, 770 con) |  |  |
|  | Spikevax | 1 |  | 66.7 (65.9 to 67.6) |  | 95.2 (91.1 to 97.4) |  |  |
|  |  | 2 |  | 95.0 (94.3 to 95.6) |  |  |  |  |
|  | Any | 1 |  | 54.1 (53.6 to 54.5) |  | 91.5 (89.9 to 92.8) |  |  |
|  |  | 2 |  | 91.0 (90.7 to 91.2) |  | 99.0 (97.7 to 99.5) |  |  |
| 40 to 64 | Comirnaty | 1 | 49.4 (45.7 to 52.9) | 45.5 (43.4 to 47.5) | 91.4 (82.4 to 95.7) | 92.5 (88.7 to 95.0) |  |  |
| from Feb 1 |  | 2 | 92.9 (87.9 to 95.8) | 80.6 (80.1 to 81.1) | 79.1 (9.5 to 95.2) | 97.9 (97.3 to 98.3) |  |  |
|  | Vaxzevria | 1 | 48.8 (46.2 to 51.4) | 34.8 (33.2 to 36.4) | 77.6 (68.4 to 84.1) | 84.0 (81.2 to 86.4) |  |  |
|  |  | 2 | 80.9 (75.2 to 85.3) | 62.8 (62.1 to 63.5) | 79.8 (29.4 to 94.2) | 94.8 (94.2 to 95.3) |  |  |
|  | Spikevax | 1 | 61.8 (7.3 to 84.2) | 56.0 (52.7 to 59.0) |  | 95.7 (86.5 to 98.6) |  |  |
|  |  | 2 |  | 93.9 (93.1 to 94.6) |  |  |  |  |
|  | Any | 1 | 49.1 (46.8 to 51.3) | 39.2 (37.8 to 40.5) | 82.5 (76 to 87.2) | 86.5 (84.3 to 88.4) |  |  |
|  |  | 2 | 85.4 (81.5 to 88.5) | 66.2 (65.5 to 66.8) | 80.8 (47.3 to 93) | 95.5 (95.0 to 95.9) |  |  |
| 65+ | Comirnaty | 1 | 54.4 (50.3 to 58.2) | 54.7 (43.2 to 63.9) | 79.3 (71.8 to 84.7) | 100.0 (no cases 1963 controls) | 74.1 (65.7 to 80.4) |  |
| from Jan 4 |  | 2 | 94.1 (90.5 to 96.4) | 67.4 (64.5 to 70.0) | 100.0 (90 to 100) | 94.6 (93.3 to 95.7) | 96.9 (91.3 to 98.9) |  |
|  | Vaxzevria | 1 | 54.4 (49.9 to 58.4) | 34.0 (22.3 to 43.9) | 79.3 (70.8 to 85.3) | 86.8 (74.3 to 93.2) | 79.9 (68.5 to 87.2) |  |
|  |  | 2 | 87.0 (81.2 to 91.0) | 49.7 (45.4 to 53.7) | 93.1 (66.4 to 98.6) | 88.7 (86.2 to 90.7) | 100.0 (no cases, 13052 controls) | |
|  | Any | 1 | 54.6 (50.9 to 57.9) | 41.4 (32.4 to 49.2) | 79.8 (73.7 to 84.5) | 91.8 (84.1 to 95.7) | 75.9 (68.6 to 81.4) |  |
|  |  | 2 | 90.6 (87.2 to 93.2) | 57.1 (53.4 to 60.5) | 96.9 (85.5 to 99.4) | 91.3 (89.5 to 92.9) | 97.4 (92.6 to 99.1) |  |
| 80+ early | Comirnaty | 1 | 53.8 (45.2 to 61.0) |  | 72.1 (57.0 to 81.9) |  |  |  |
| d1 Dec8-Jan 3 |  | 2 | 81.0 (75.6 to 85.1) | 42.9 (7.9 to 64.7) | 93.2 (84.0 to 97.1) | 73.4 (53.2 to 84.8) |  |  |
| *d1: 28 days after first dose to time of second (if given), d2: 14 days after second dose | | | | | | | |  |
