## Supplementary figures and images for "Vaccine effectiveness and duration of protection of Comirnaty, Vaxzevria and Spikevax against mild and severe COVID-19 in the UK"

### Supplementary Figure S1

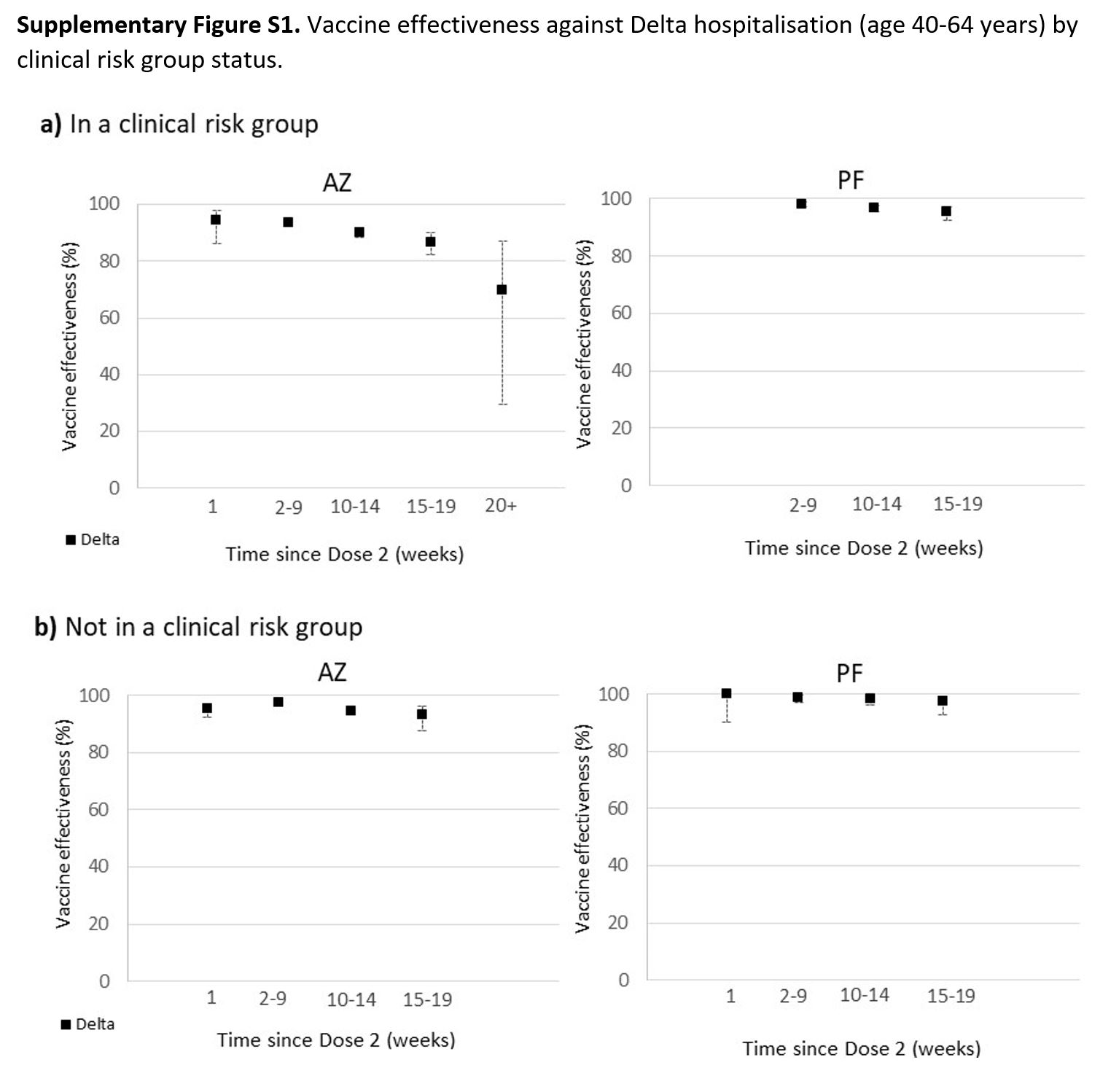

### Supplementary Figure S2

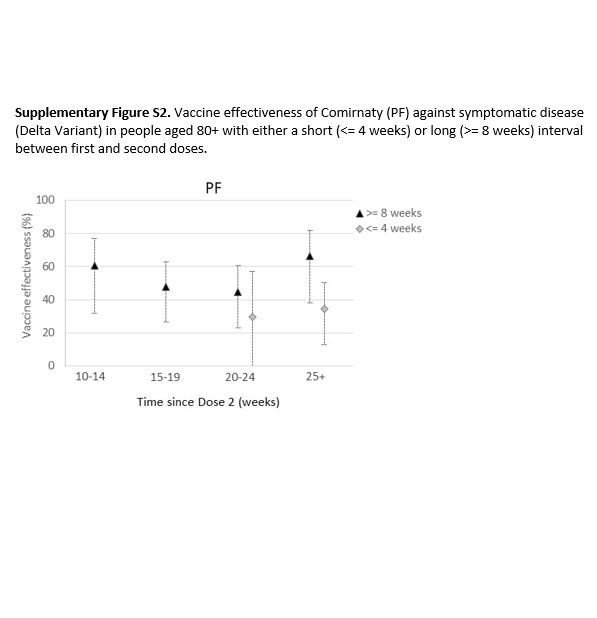

### Supplementary Figure S3

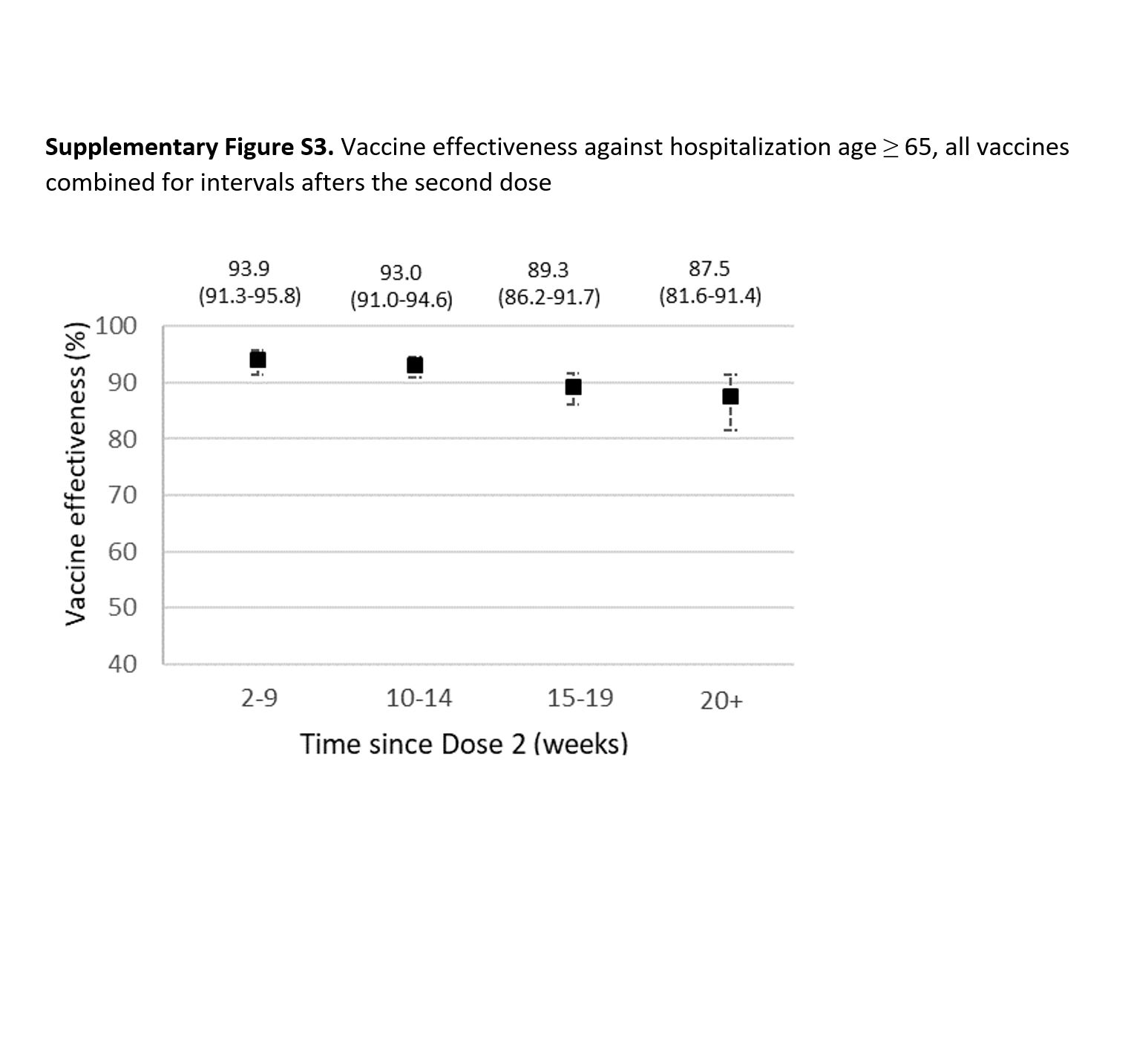
